# Comparative Clinical Evaluation of the Alinity m STI Multiplex PCR Assay for Diagnosis and Surveillance of *Chlamydia trachomatis*, *Neisseria gonorrhea*, *Trichomonas vaginalis*, and *Mycoplasma genitalium*

**DOI:** 10.1101/2023.12.04.23299219

**Authors:** Amorce Lima, Dominic Uy, Joshua Kostera, Suzane Silbert

## Abstract

**Background:** *Chlamydia trachomatis* (CT) and *Neisseria gonorrhoeae* (NG) are routinely tested and reported; however, *Trichomonas vaginalis* (TV) is the most common STI in the US and the prevalence of *Mycoplasma genitalium* (MG) infections is likely higher than estimated. We examined the clinical performance of the Alinity m STI assay for detection and surveillance of CT/NG/TV/MG in urine specimens from patients at a large academic medical center.

**Methods:** Prevalence of mono- and co-infections on Alinity m STI pathogens and predictors of a positive result were identified. Alinity m STI and Aptima Combo 2 CT/NG and TV assay (Panther System) results were compared, with discrepant results run on the cobas 6800 CT/NG, TV and MG assays. Analyzer turnaround times (TAT) were determined for Alinity m and Panther systems.

**Results:** 199 urine specimens were included. Age ≥25 years, collection outside the emergency department (ED), and asymptomatic status were predictive of TV or MG infection; symptomatic status was the only predictor of CT or NG infection. Overall agreement rates for the Alinity m, Aptima, and cobas assays ranged from 86.4% to 99.5% for the four pathogens. TV and MG infections comprised 54% of the positive samples and were more often asymptomatic than CT and NG infections. Analyzer TAT (onboard to result reporting) was 4 hours 45 minutes for the Aptima CT/NG, 3 hours 25 minutes for Aptima TV, and 1 hour 55 minutes for Alinity m STI assay.

**Conclusions:** The Alinity m STI assay allows for fast and simultaneous detection of the four major STI pathogens, which can facilitate surveillance and provide accurate results to help clinicians diagnose for initiation of appropriate treatment.

## Introduction

The most recent CDC surveillance data indicates that 2.5 million STIs were reported in 2021, an increase of almost 3% from 2020 (1). The four most common STI pathogens are *Chlamydia trachomatis* (CT), *Neisseria gonorrhoeae* (NG), *Trichomonas vaginalis* (TV), and *Mycoplasma genitalium* (MG). Of these, TV is the most common, with 1.2% prevalence among 2013-2014 NHANES participants (2); however, TV is not considered reportable by CDC (3, 4). Current guidelines recommend TV screening only in high-risk populations (3). As the majority of individuals with TV infection are asymptomatic, it can remain undiagnosed and untreated for years, leading to severe consequential health outcomes such as pelvic inflammatory disease, urethritis, and infertility. MG has emerged as an STI pathogen of concern with drug resistant MG placed on CDC’s Watch List in 2019 (5). Yet MG is not classified as reportable by CDC; current guidelines recommended MG testing in only a small subset of patients with recurrent urethritis or cervicitis (3). Thus, the true prevalence of MG infection is not clear and is likely higher than expected (6).

Untreated STIs have significant long-term health consequences and inflict a substantial economic medical burden (7). Additionally, NG, TV, and MG infections are associated with increased risk for acquiring or transmitting HIV (3, 4, 8). Timely diagnosis of STIs is essential to reduce transmission and initiate treatment (3), but diagnosis based on patient symptoms alone can be challenging. Co-infections by more than one pathogen can cause overlapping symptoms that confound both diagnosis and initiation of appropriate therapy. Inaccurate diagnosis leading to inappropriate or inconsistent treatment can lead to the emergence of multidrug resistant strains (9–11).

Guidelines recommend screening for the reportable pathogens, CT and NG, with nucleic acid amplification tests (NAATs) to help reduce transmission and guide treatment (12, 13). Similar NAAT-based surveillance programs for TV and MG would be expected to identify subclinical or asymptomatic infections, refine treatment algorithms, and reduce transmission rates (3, 14, 15). Because sequential testing for each pathogen can prolong the time to diagnosis and delay initiation of appropriate treatment, NAATs have been developed for simultaneous detection of multiple pathogens from a single specimen.

The Alinity m STI assay (Abbott Laboratories, Des Plaines, IL, USA) is an in vitro multiplex RT-PCR assay that is designed for qualitative and simultaneous detection of CT, NG, MG, and TV in various urogenital and extragenital specimens. The objective of this study was to evaluate the Alinity m STI assay and compare the results with our standard of care (SOC) STI tests, the Aptima Combo 2 CT/NG and the Aptima TV assays, run on the Panther system (Hologic, Marlborough, MA) for urine specimens collected from patients at a large academic medical center in the United States. We also examined the utility of the Alinity m STI assay for surveillance of TV and MG infection.

## Materials and Methods

### Specimens and setting

199 residual de-identified urine specimens from consecutive inpatients, outpatients, and patients in the emergency department (ED) at Tampa General Hospital (TGH; Tampa, Florida, USA) were included in the study after SOC testing. No inclusion or exclusion criteria were applied. The study was conducted following GCLP and in accordance with the Declaration of Helsinki, under IRB# 1301043.

### Specimen collection and disposition

Approximately 2 mL of first catch urine specimen was transferred from the primary urine collection container to an Aptima Urine Specimen Transport Tube for SOC testing on the Panther system (Figure 1). Two additional 5-mL aliquots from the primary urine collection container were pipetted into cryogenic vials: Aliquot #1 was transferred to the Alinity m multi-collect tube which was used for testing on the Alinity m STI assay and Aliquot #2 was used for referral testing to confirm MG positives on Alinity m and resolve any discrepant results.

**Figure 1.**
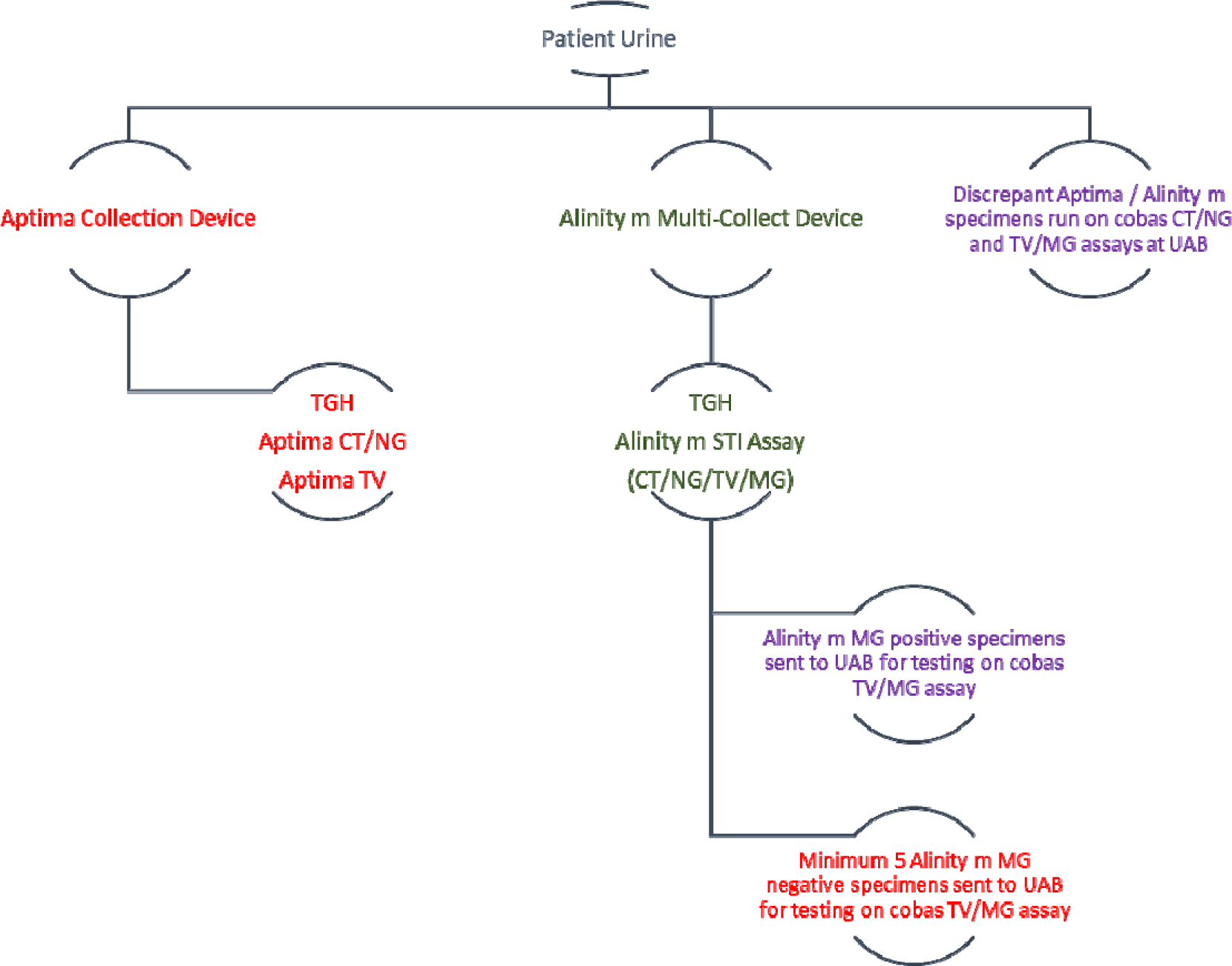
Study design.

All MG-positive specimens and at least 5 MG-negative specimens on Alinity m STI with sufficient remaining volume were sent to the University of Alabama STD Diagnostics Lab (UAB; Birmingham, AL, USA), to be tested for CT, NG, TV, and MG on the cobas 6800 system (Roche, Branchburg, NH, USA). All specimens with discrepant results on the Aptima and Alinity m assays with sufficient volume for Aliquot #2 were also sent to UAB for testing with the cobas 6800 CT/NG, TV and MG assays.

### Assay platforms

The Alinity m STI assay (Abbott Laboratories) was run on a single Alinity m analyzer at TGH according to the manufacturer’s instructions using reagents that was not yet approved by the FDA for that purpose. The Alinity m STI assay primers and probes amplify and detect an endogenous human DNA sequence as validity control (cellular control, CC) and an exogenous internal control (IC) is used to confirm the absence of PCR inhibitors in the test specimen. For this study, 2.1 mL of each collected urine specimen was transferred from Aliquot #1 to an Alinity m multi-Collect tube for testing and the analyzer was programmed to test all four STI pathogens (CT, NG, MG, and TV). Alinity m reagents were left onboard the analyzer for 10-30 days.

The Aptima Combo 2 for CT/NG and Aptima TV assays were run on the Panther system (Hologic, Inc.) at TGH. If clinical orders were for CT/NG testing only, then TV testing was performed using the same specimen. Because onboard stability of the Aptima reagents is 72 hours, TV testing was performed in batches. Upon completion of testing, the reagents were removed from the Panther instrument and refrigerated. If TV testing was not done on the same day as CT/NG testing, specimens were stored at 2-8°C until the next batch run.

The cobas 6800 CT/NG assay and cobas 6800 TV/MG assays were run at UAB. All assays were run following the manufacturers’ instructions.

### Workflow study

Total TAT for the Alinity m STI assay were calculated based on timepoints entered into the laboratory information system and electronic medical record. The average time from specimen receipt to results reporting and from specimen loading onto the analyzer to results reporting were tracked.

### Data analysis

The prevalence of each analyte and co-infections in the study population were determined based on the Alinity m STI assay results. Positive, negative, and overall percent agreement (PPA, NPA, OPA) and 95% confidence intervals (CIs) and kappa values were determined for results from the Alinity m STI assay and the comparator assays. Odds ratios and 95% CIs were calculated to identify variables predictive of positive Alinity m STI assay results.

## Results

### Study cohort characteristics and test results

Urine specimens from 199 patients were included in the study; demographic characteristics are shown in Table 1. The majority of specimens were collected from females and in the inpatient setting. Median patient age was 29 years; of note, a patient between the age of 6-10 was included, but was the only patient under 16 years of age and was negative for all pathogens tested on the Aptima CT/NG and TV assays and Alinity m STI assay (CT/NG/TV/MG). The cohort was diverse, with slightly more Black patients than White or Hispanic patients. Eighteen patients in the “Other” category were Asian, mixed-race, Middle Eastern, or chose not to identify their racial/ethnic background.

**Table 1.**
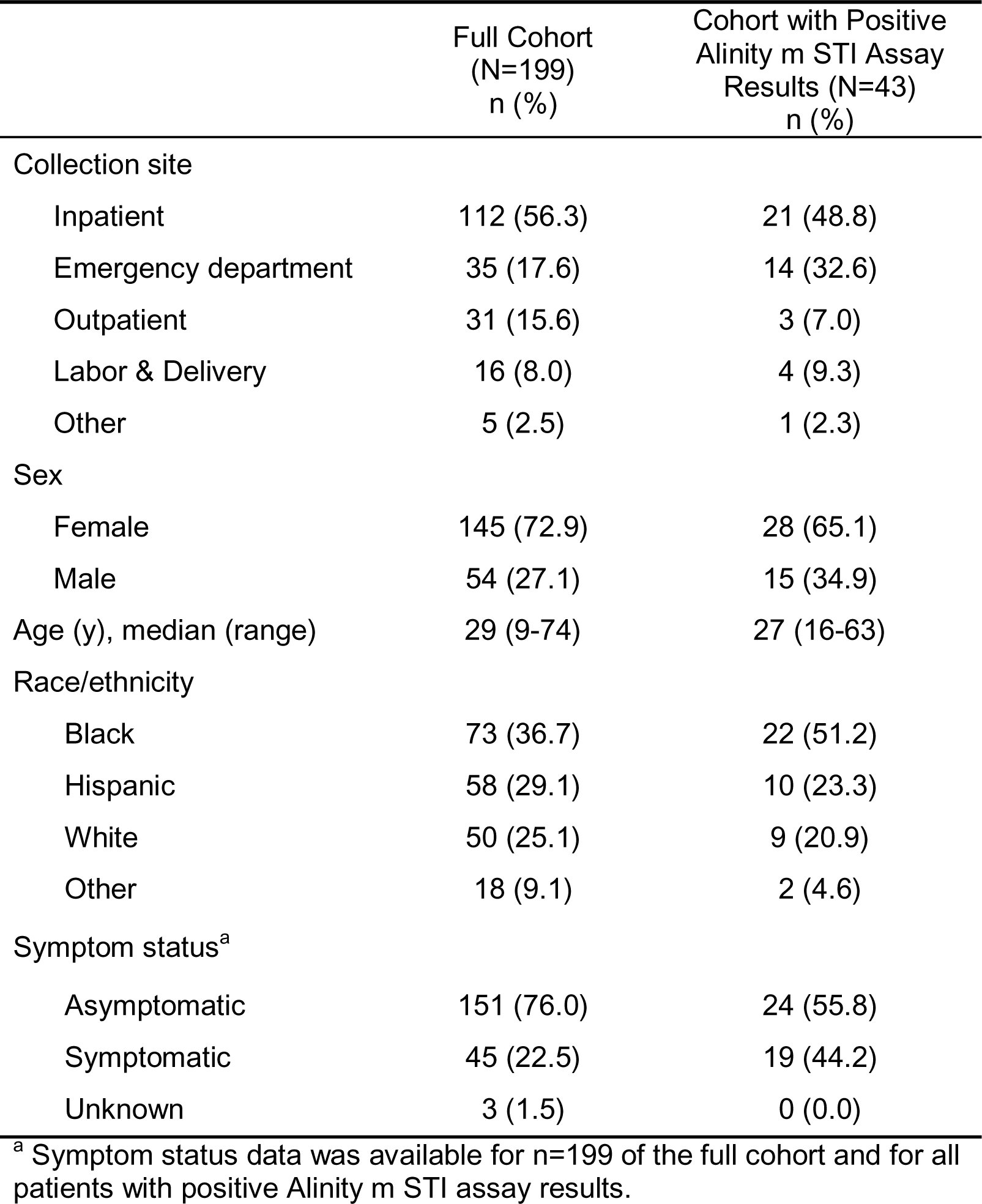
Study Cohort Demographics.

Forty-three patients had positive Alinity m assay results (Table 1). There were 7 CT, 4 NG, 12 TV, and 11 MG mono-infections; 3 CT/NG co-infections; 3 CT/MG co-infections; 2 NG/TV co-infections; and 1 TV/MG co-infection. No triple or quadruple co-infections were detected. TV and MG infections comprised more than half (54%) of the positive samples and 14% were co-infections with CT or NG (Figure 2A). Only 25% of positive results were CT or NG mono-infections and 7% were CT/NG co-infections. The mean age of patients with TV or MG mono-infections or TV/MG co-infection was 32 years, 5 years older than the mean age of patients with CT or NG mono-infection or CT/NG co-infection. In this cohort, TV, MG, TV/MG, and associated co-infections were mostly asymptomatic (20 asymptomatic and 9 symptomatic), whereas CT, NG, CT/NG infections were more often symptomatic (10 symptomatic and 4 asymptomatic). The distribution of Alinity m STI positive results varied based on site of specimen collection (Figure 2B). The vast majority of positive specimens collected in the inpatient setting were from asymptomatic patients, whereas the majority of positive specimens collected in the ED and all positive specimens collected in the outpatient setting were from symptomatic patients. Specimens were tested on 82 unique testing days over the course of 10 months.

**Figure 2.**
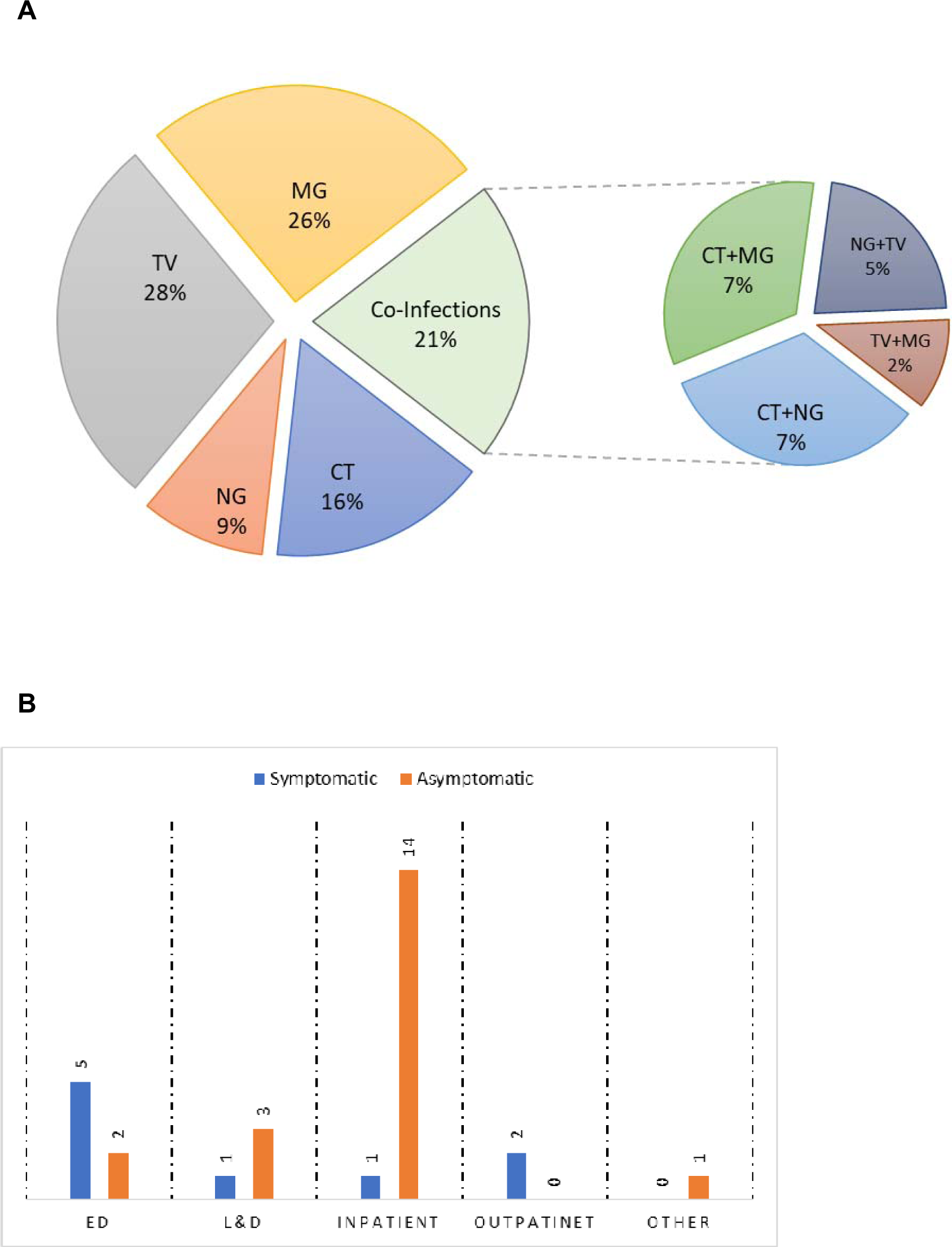
Alinity m STI assay positive specimens in the study cohort. (A) Distribution by mono-infection or co-infection. (B) Number of positive specimens collected in each setting from asymptomatic or symptomatic patients.

### Predictors of positive Alinity m STI assay results

Factors that were most predictive of an assay result positive for TV or MG mono- or co-infection were age ≥25 years, specimen collected at a site outside the ED, and asymptomatic status (Figure 3A). The only factor predictive of CT, NG, or CT/NG positive results was symptomatic status (Figure 3B). Specimen collection outside the ED was somewhat predictive of a positive CT, NG, or CT/NG result.

**Figure 3.**
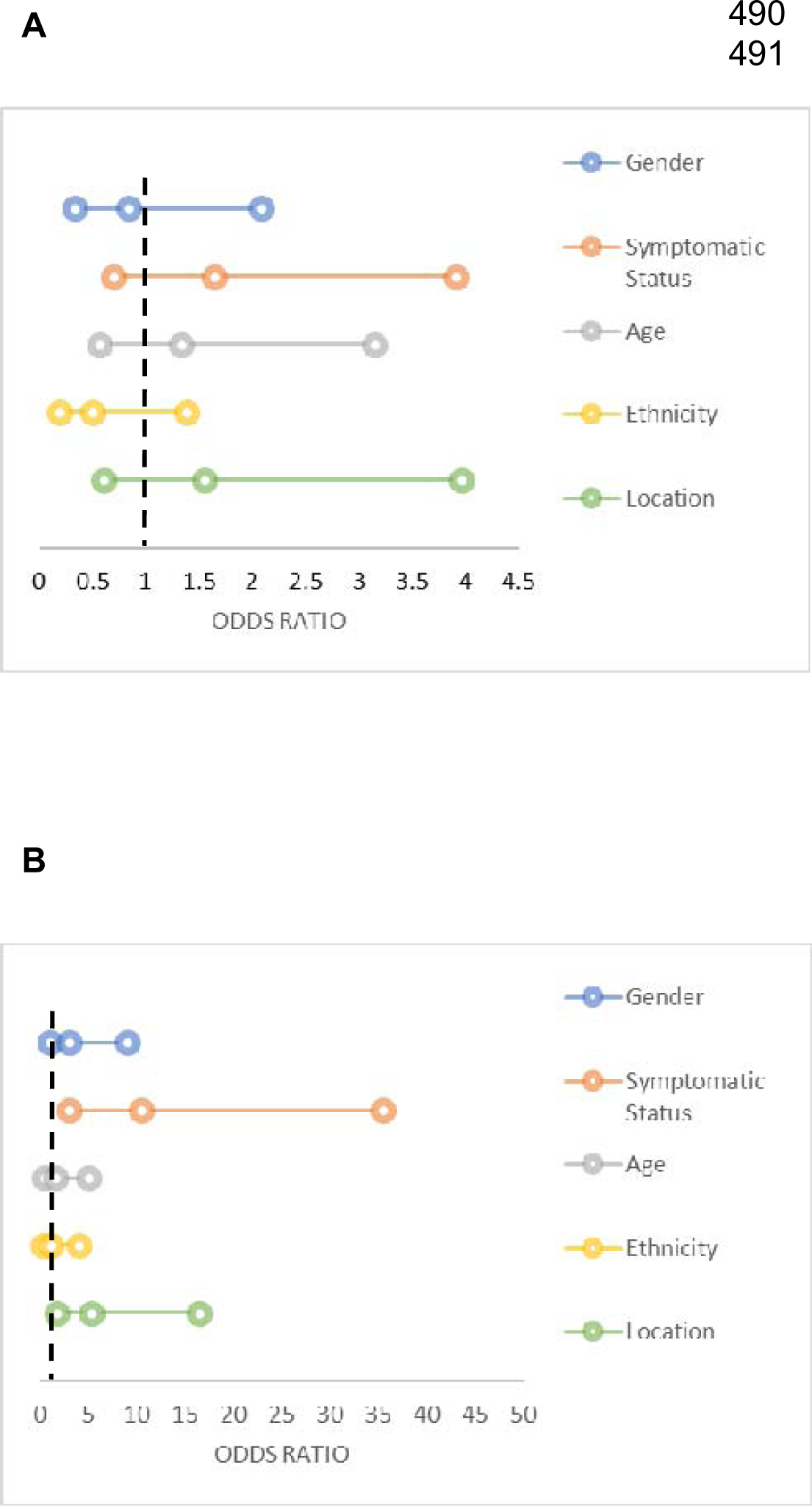
Factors predictive of an Alinity m STI assay positive result. Odds ratios and 95% confidence intervals for factors predicting (A) a positve TV, MG, TV/MG, TV/NG, or MG/CT result or (B) a postive CT, NG, or CT/NG result on the Alintiy m STI assay.

### Comparative performance of STI NAATs

Agreement between Alinity m STI, Aptima CT/NG, Aptima TV, and cobas MG assay results is summarized in Table 2. Overall percent agreement (OPA) between Alinity m STI and comparator assay results was 99.5% (95%CI: 97.2%, 99.9%) for CT, 99.5% (95%CI: 97.2%, 99.9%) for NG, 98.4% (95%CI: 95.5%, 99.5%) for TV, and 86.4% (95%CI: 66.7%, 95.3%) for MG.

**Table 2.**
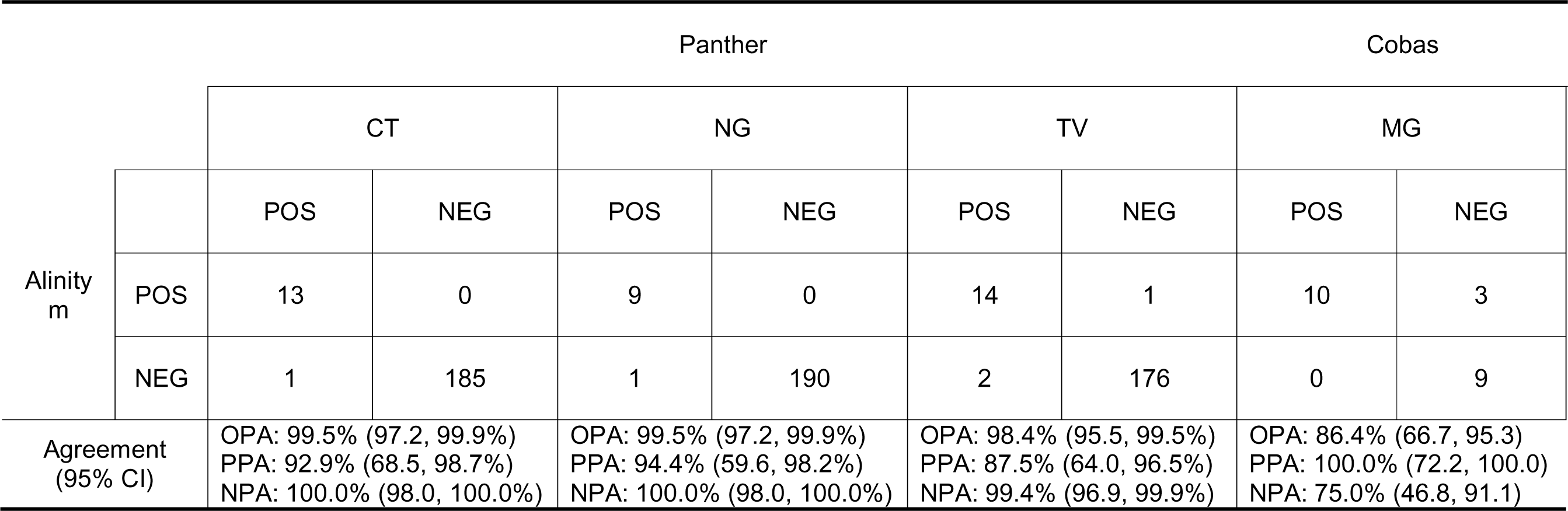
Agreement Between Alinity m STI Assay (CT/NG/TV/MG) and Aptima Combo 2 & Trichomonas vaginalis Assays (CT/NG TV) or Cobas TV/MG Assay (MG) Results.

For CT, 1 specimen collected from an asymptomatic female patient in her 20s was CT-positive on Aptima, CT-negative on Alinity m, and CT-positive on cobas. The review of the Aptima CT/NG relative light units (RLU) for this specimen was 829 vs mean RLU for all CT-positive specimens was 1125 (SD 227), indicating a low CT concentration in the specimen that was not detected by Alinity m STI.

One specimen collected from an asymptomatic female patient under 20 years old was NG-positive on Aptima and NG-negative on Alinity m. The RLU on Aptima CT/NG for this specimen was 563, more than 3 SD from the mean RLU for all NG-positive specimens of 1299 (SD 154). The specimen was also found to be NG-negative on cobas, in agreement with the Alinity m STI result.

Two specimens were found to be TV-positive on Aptima and TV-negative on Alinity m STI assay. The first was from an asymptomatic male patient in his 50s that was TV-negative on cobas. The specimen was not positive for any other pathogen. The specimen RLU on Aptima TV was 133, more than 2 SD from the mean RLU of 1324 (SD 369) for all TV-positive specimens from asymptomatic patients. The second specimen, from an asymptomatic female patient in her early 40s, was TV-positive on cobas. The TV RLU was 1528, within 1 SD of the mean RLU of 1324. One specimen from an asymptomatic female in her 30s was TV-negative on Aptima, TV-positive on Alinity m, and TV-negative on cobas.

Three specimens were MG-negative on cobas but MG-positive on Alinity m. One specimen from a symptomatic female patient and another from an asymptomatic female patient in their 20s were negative for CT/NG/TV on Alinity m, negative for CT/NG and TV on Aptima, and negative for CT/NG and TV/MG on cobas. The third specimen, from an asymptomatic female patient in her 30s, was also TV-positive on the Alinity m, Aptima, and cobas assays. The cause for the discrepancy between Alinity m and cobas MG results could potentially be attributable to neat sample degradation between collection and testing with cobas or assay sample type sensitivities.

For the 8 specimens with discrepant results on the Aptima, Alinity m, and cobas assays, there was no significant difference in the Alinity m IC and CC cycle number (CN) values compared to the average IC and CC CN values of all specimens in cohort (see Table, Supplemental Digital Content 1).

### Aptima and Alinity m STI assay performance by symptom status

Although the assays are qualitative, we compared assay readouts (RLU for Aptima and CN for Alinity m) across target pathogens and in specimens from symptomatic and asymptomatic patients. Aptima RLUs were significantly higher for specimens from symptomatic patients with CT/NG co-infection compared to those with CT, NG, or TV mono-infection or in asymptomatic patients (Figure 4A). It is not surprising that higher RLUs were seen with co-infections, as they would elicit a double probe response. CN values for specimens run on the Alinity m STI assay were consistently between 10 and 40 and did not differ by symptom status (Figure 4B).

**Figure 4.**
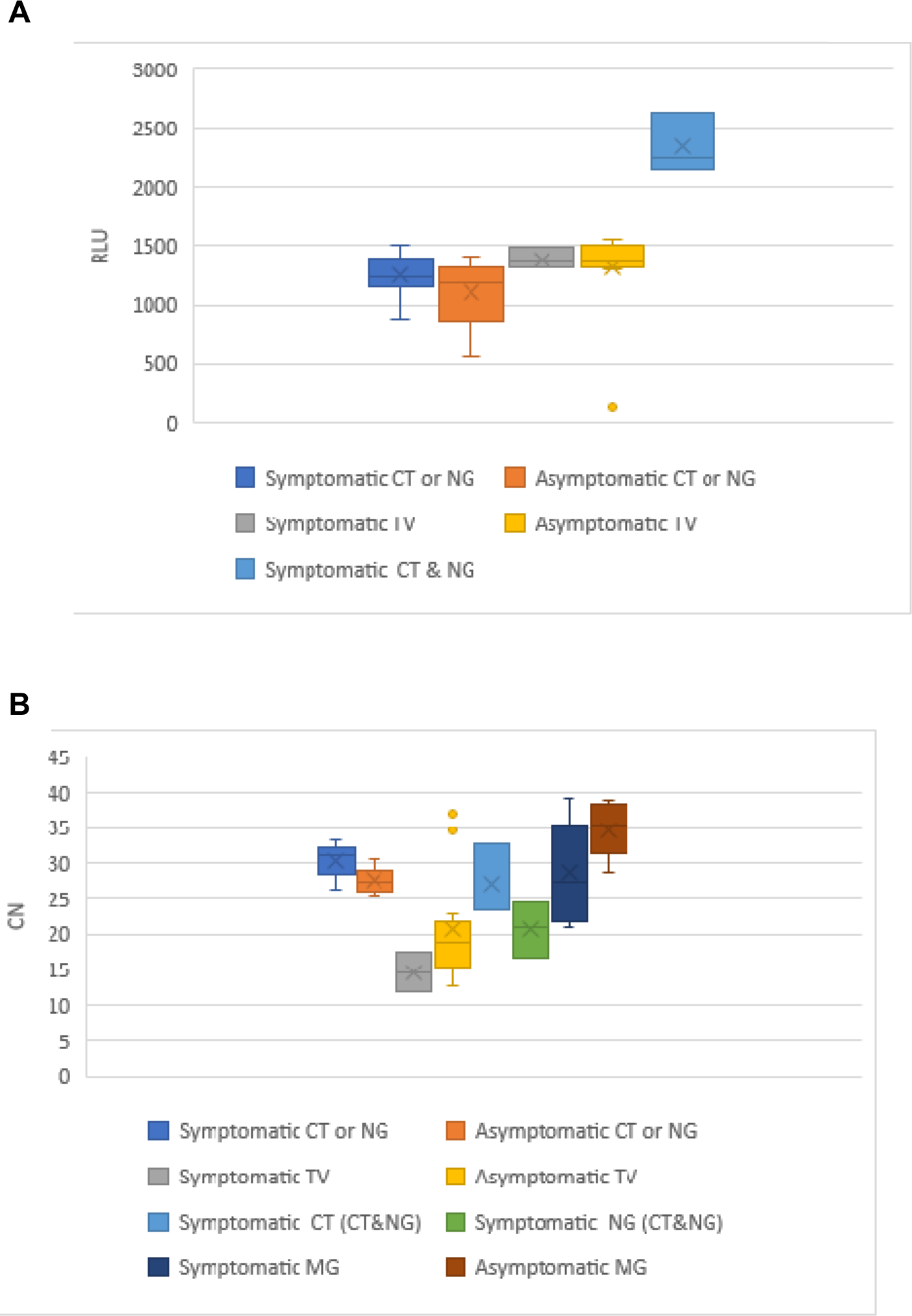
(A) Aptima Combo 2 RLU values and (B) Alinity m STI assay CN values for positive specimens from symptomatic and asymptomatic patients.

### Aptima and Alinity m STI workflow analysis

We compared the workflows for each assay for 199 specimens. The average overall turnaround time, from receipt of the specimen in the lab to results reporting, for the Alinity m STI assay for all four analytes was 6 hours 59 minutes (n=199; Figure 5A). The overall turnaround time was 8 hours 28 minutes (n=199) for the Aptima CT/NG assay and 80 hours 56 minutes (n=128) for the Aptima TV assay, reflecting the batching of specimens. To assess workflow independent of batching, we examined the turnaround time from loading the specimen on the analyzer to results reporting for the three assays (Figure 5B). The average time from onboarding to result was 3 hours 25 minutes for the Aptima CT/NG assay, 3 hours 25 minutes for the Aptima TV assay, and 1 hour 55 minutes for the Alinity m STI assay for CT/NG/TV/MG.

**Figure 5.**
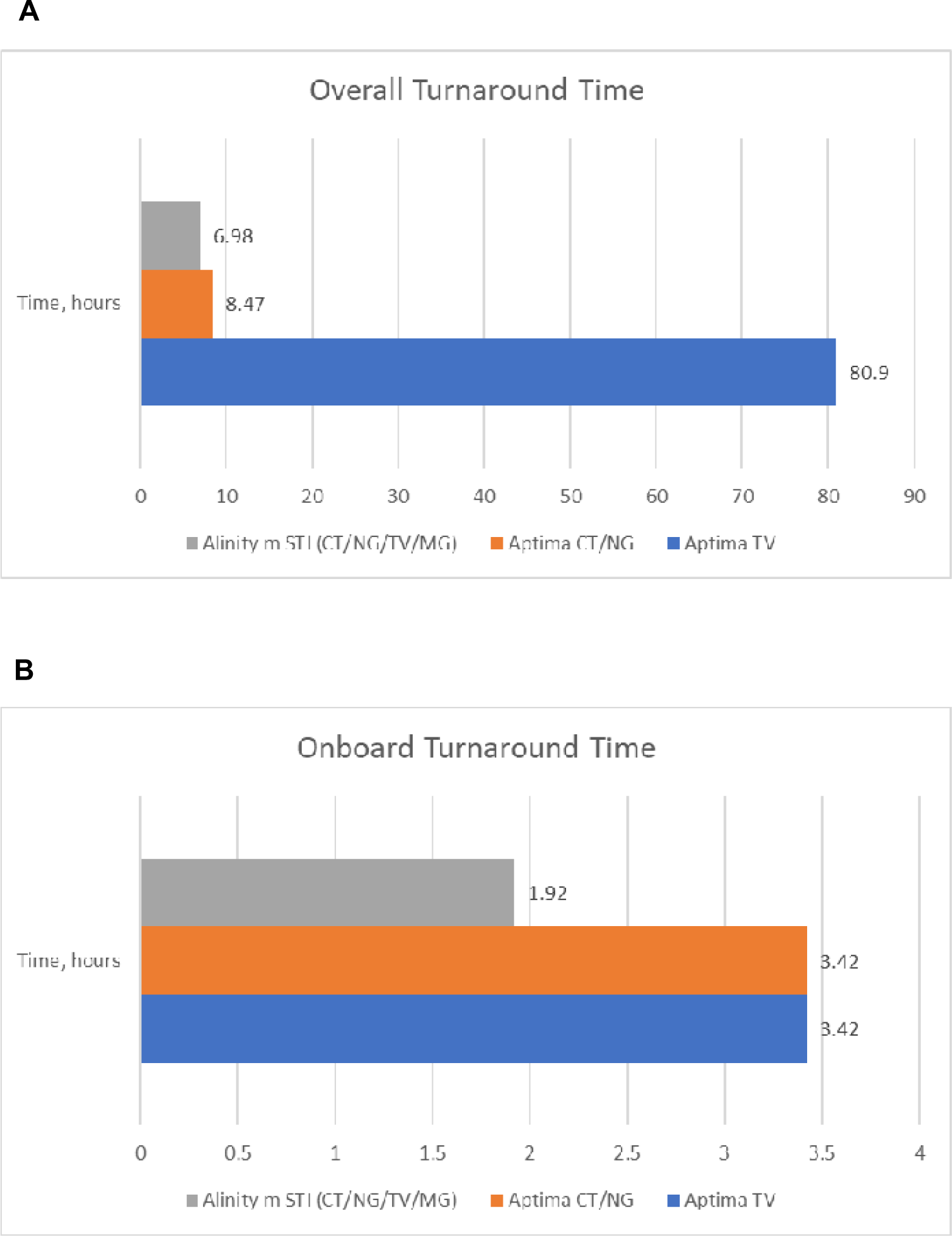
Turnaround times for Alinity m STI assay on the Alinity m platform compared to Aptima CT/NG and TV assays run on the Panther platform. (A) Overall turnaround from specimen arrival in the lab to results reporting. (B) Onboard turnaround from specimen loading on the analyzer to results reporting.

## Discussion

In this study, we found high agreement rates between the Alinity m STI assay and the Aptima Combo 2 and TV assays for CT/NG and TV (OPA ≥98.4%) or cobas TV/MG assay for MG (OPA=86.4%). Discrepant specimens that were negative on Alinity m were primarily seen in asymptomatic patients where a CN value was not generated or had high CN values greater than the assay cutoff cycle, indicating low levels of the pathogen target. These findings are consistent with a previous study demonstrating concordance of results from the Alinity m STI assay and RealTime CT/NG/TV assay run on the m2000 system (16).

The prevalences of infection and co-infection in our study indicate that TV and MG infections are likely being missed. Only 2 out of the 14 positive TV samples had a microscopy test performed and both were negative. Current practice at our institution is consistent with CDC guidance for CT and NG screening (3), with specimens run on the Aptima CT/NG assay as the SOC test. We do not routinely test for TV, unless ordered by the provider and do not test for MG. In our cohort, 68% of positive samples on Alinity m were TV or MG positive (53% mono-infection) and were more likely to be from asymptomatic patients.

Our findings are consistent with the literature that most TV/MG infections are asymptomatic. The Swiss STAR trial screened high-risk women and found a prevalence of undiagnosed TV up to 10.4% and MG up to 6.7% (17). A US study of 374 female adolescents in juvenile detention found that 8% tested positive for TV and half of these were asymptomatic (18). Another study of men who have sex with men (MSM) with HIV who were screened for MG found a 20% prevalence, 93% of which were asymptomatic (19). Addition of TV and MG testing would reduce the likelihood of missed infections, decrease the risk of transmission, and improve appropriate antimicrobial use.

Identification of factors predictive of positive TV and MG results on Alinity m STI or other NAATs could help inform adjustments to STI screening algorithms (1, 3) to ensure that asymptomatic infections are detected and treated. In our study, the mean age of patients with TV/MG-positive results was 5 years older than those with CT/NG-positive results. This is consistent with several surveillance studies showing that TV/MG infections are more likely to be diagnosed at a later age than CT/NG infections (3, 20–22). A recent US study of NHANES data found that women, smokers, non-Hispanic Black patients, and patients of lower socioeconomic status are more likely to have a TV infection (23). In contrast to our findings, a study of a high-risk population of women in New Mexico found that MG infection was more prevalent in younger than older women (24). Rigorous, nationwide studies of predictors of MG infection are needed to develop evidence-based recommendations for MG testing.

The distribution of positive Alinity m results by collection site illustrated that the ED is utilized to confirm infection in symptomatic patients. Studies suggest that a growing number of STIs are being diagnosed in the ED setting, possibly related to closure of STI clinics and increased utilization of the ED for primary care (25, 26). STI testing in the ED may help identify individuals with asymptomatic infection, allowing earlier initiation of treatment to stop transmission chains. A recent study developed a prediction model for identifying patients as candidates for CT/NG/TV testing in the ED, based on age, marital status, race, findings from vaginal wet prep, and urinalysis (27).

In the workflow analysis, Alinity m STI reduced turnaround times compared to Aptima by removing the need for batching of TV assays. Simultaneous testing of all four pathogens also decreased the onboard to result turnaround times. Our findings are consistent with a previous report of faster turnaround times with the Alinity m STI assay compared to the Abbott RealTime assay run on the m2000 instrument (28). A workflow study of the Alinity m platform also demonstrated reduced the time to diagnosis and treatment initiation (29). More rapid results reporting has implications for management of STIs in the ED setting. A recent study suggested that rapid NAAT for STIs may help direct appropriate utilization of antimicrobials in the ED and reduce the risk of treatment resistance (30).

This was a single-center study, which may limit the generalizability of our findings; however, we utilized a third assay system run at a separate facility to resolve discrepancies in assay results. The Aptima system was not used to test for MG, which prevented a direct comparison to the Alinity m STI assay. This study also only utilized urine specimens; additional studies are needed to evaluate the performance of the Alinity m STI assay with other genital and extragenital specimen types and collection methods. Despite these limitations, this study utilized a convenience sample with no inclusion or exclusion criteria, which led to a diverse study cohort. We collected specimens from multiple sites in the medical center, and tested throughout the year to ensure that the sample was not enriched for certain peak periods.

Our findings support the surveillance of MG and TV, which often remain as asymptomatic infections that serve as ongoing transmission reservoirs. Symptoms alone are non-diagnostic and can overlap significantly between CT, NG, TV, and MG. The Alinity m STI assay allows for simultaneous detection of the four major STI pathogens in a single assay, which can simplify the implementation of MG/TV surveillance programs in health systems that already report NG and CT infections. TV and MG surveillance programs can inform evidence-driven changes in the STI diagnostic algorithm, with multiplex assays moving patients more quickly to diagnosis and initiation of the most appropriate treatment.

## Data Availability

All pertinent data produced in the study are included in the manuscript.

## Acknowledgements

The authors would like to thank Yan Zhang, PhD, for assistance with the statistical analysis. Stacey Tobin, PhD, provided editorial support for manuscript preparation, with compensation from Abbott Laboratories.

**List of Supplemental Digital Content**

**Supplemental Digital Content 1.pdf**

