## Supplemental Table 1 for "Comparative Clinical Evaluation of the Alinity m STI Multiplex PCR Assay for Diagnosis and Surveillance of *Chlamydia trachomatis*, *Neisseria gonorrhea*, *Trichomonas vaginalis*, and *Mycoplasma genitalium*"

**Supplemental Digital Content 1.** Cycle Number (CN) Values for Alinity m STI Assay Internal Control (IC) and Cellular Control (CC) for Specimens with Discrepant Results^a,b^

| Analyte | Discrepant Result | IC | CC |
| --- | --- | --- | --- |
| CT | Aptima POS / Alinity m NEG | 30.40 | 29.13 |
| NG | Aptima POS / Alinity m NEG | 31.02 | 30.41 |
| TV | Aptima POS / Alinity m NEG | 30.82 | 32.90 |
|  | Aptima POS / Alinity m NEG | 29.29 | 27.16 |
|  | Aptima NEG / Alinity m POS | 29.93 | 28.27 |
| MG | Cobas NEG / Alinity m POS | 29.84 | 27.67 |
|  | Cobas NEG / Alinity m POS | 30.43 | 26.10 |
|  | Cobas NEG / Alinity m POS | 31.02 | 28.35 |
| ^a^ Mean±SD CN values for the full cohort were IC=30.56±0.73, CC=29.61±3.02.  ^b^ Mean±SD CN values for specimens positive for any analyte were IC=30.08±0.76 and CC=27.88±2.77. | | | |
